# Forecast verifications for the real COVID-19 pandemic dynamics in Qatar

**DOI:** 10.1101/2021.06.16.21259018

**Authors:** Igor Nesteruk

## Abstract

The COVID-19 pandemic dynamics in Qatar in the second half of May and the first half of June 2021 was compared with the published results of SIR-simulations based on the data from the period April 25 - May 8, 2021. Forecast verification showed very good agreement with the real number of cases (which can exceed the laboratory-confirmed one more than 5 times). The positive effect of mass vaccination became visible in June 2021.

## Introduction

The COVID-19 pandemic dynamics in Qatar was simulated with the use of SEIR-model (susceptible-exposed-infected-removed) [1] and SEIRD-model (susceptible-exposed-infected-removed-dead) [2]. Some recent SIR-simulations [3, 4] were based on the datasets about the number of cases in December 2020 and April 25 – May 8, 2021 available in COVID-19 Data Repository by the Center for Systems Science and Engineering (CSSE) at Johns Hopkins University (JHU), [5]. In particular, the incompleteness of the statistical information has been taken into account in [4] with the use of method proposed in [6]. It was shown that the actual accumulated number of COVID-19 cases may exceed the laboratory-confirmed number of cases registered in Qatar more than 5 times. Comparison with the corresponding SIR-curves has showed that the effect of mass vaccination was not evident during 4 months after its onset in December 2020, [4]. In this study, we will follow the epidemic in Qatar until mid-June 2021 in order to verify the predictions and to investigate the impact of vaccination.

### Data and monitoring changes in epidemic dynamics

We will use tree data sets regarding the accumulated numbers of confirmed COVID-19 cases *V*_*j*_, number of vaccinated people *P*_*j*_ and number of vaccinations *Q*_*j*_ in Qatar from JHU, [5]. These values and corresponding moments of time *t*_*j*_ (measured in days) are shown in Table 1. For SIR-simulations, the values of *V*_*j*_ and *t*_*j*_ corresponding to the time period: April 25 – May 8, 2021 were used in [4]. The values presented in Table 1 and datasets available in [4] were used only for monitoring the changes in epidemic dynamics, which can be caused by vaccination, changes in quarantine restrictions, emergence of new strains of the pathogen, etc. To control these changes and to identify the epidemic waves, we can use daily or weekly numbers of new cases and their derivatives (see, e.g., [7-9]). Since these values are random, we need some smoothing (especially for daily amounts, which are also characterized by some weekly periodicity). For example, we can use the smoothed daily number of accumulated cases:

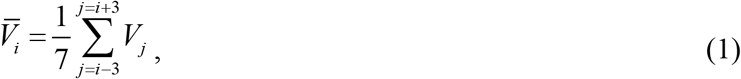

**Table 1.** Cumulative numbers for Qatar from [5]. The laboratory-confirmed Covid-19 cases *V*_*j*_, number of vaccinations *Q*_*j*_, the number of vaccinated people *P*_*j*_, and corresponding moments of time.

| Day in May<br>and June<br>2021 | Number<br>of<br>cases,<br>$V_j$ | Number<br>of vacci-<br>nations,<br>$Q_j$ | Number of<br>vaccinated<br>people<br>$P_j$ |
| --- | --- | --- | --- |
| 21 | 214830 | 2233616 | 1288584 |
| 22 | 215160 | - | - |
| 23 | 215443 | 2293240 | 1321233 |
| 24 | 215742 | 2329338 | 1341921 |
| 25 | 216091 | 2365647 | 1361511 |
| 26 | 216397 | 2403165 | 1381456 |
| 27 | 216683 | 2440930 | 1400720 |
| 28 | 216885 | 2475837 | 1420090 |
| 29 | 217041 | 2491638 | 1430121 |
| 30 | 217230 | 2514585 | 1443882 |
| 31 | 217458 | 2545193 | 1461871 |
| 1 | 217688 | 2574692 | 1479717 |
| 2 | 217882 | 2600344 | 1494265 |
| 3 | 218080 | 2622285 | 1510561 |
| 4 | 218263 | 2643573 | 1526103 |
| 5 | 218455 | 2656695 | 1531477 |
| 6 | 218627 | 2676239 | 1541863 |
| 7 | 218798 | 2700942 | 1554740 |
| 8 | 218980 | 2716670 | 1561481 |
| 9 | 219138 | 2729437 | 1566366 |
| 10 | 219281 | 2748452 | 1572212 |
| 11 | 219466 | 2768028 | 1578669 |
| 12 | 219613 | 2780916 | 1583143 |
| 13 | 219730 | 2795091 | 1587414 |

The first and second derivatives can be estimated with the use of following formulas:

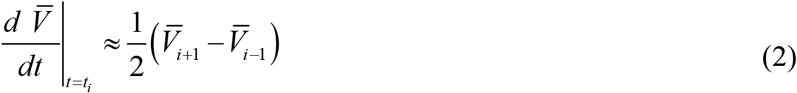

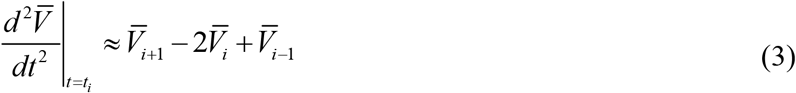

### Generalized SIR model and parameter identification procedure

In [3, 4] the generalized SIR-model and the exact solution of the set of non-linear differential equations relating the number of susceptible *S*, infectious *I* and removed persons *R* were used in order to simulate different epidemic waves (see, e.g., [6, 10]). This solution uses the function

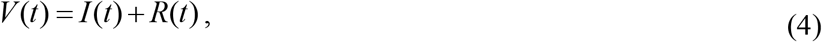

corresponding to the number of victims or the cumulative confirmed number of cases. Its derivative:

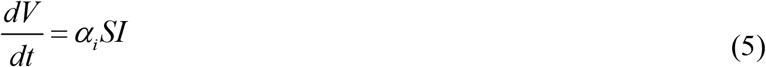

yields the estimation of the average daily number of new cases. When the registered number of victims *V*_*j*_ is a random realization of its theoretical dependence (4), the exact solution presented in [6, 10] depends on five parameters (*α*_*i*_ is one of them, *i* is the number of the epidemic wave). The details of the optimization procedure for their identification can be found in [6, 11].

If we assume, that data set *V*_*j*_ is incomplete and there is a constant coefficient *β*_*i*_ ≥ 1, relating the registered and real number of cases during the *i-th* epidemic wave:

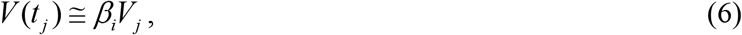

the number of unknown parameters increases by one. The values *V*_*j*_, corresponding to the moments of time *t*_*j*_ from the period April 25 – May 8, 2021 have been used in [4] to find the optimal values of these parameters corresponding to the third epidemic wave in Qatar. In particular, the optimal value of the visibility coefficient *β*_3_ = 5.308 was calculated.

## Results and Discussion

The results of SIR simulations performed in [4] are shown in the figure. Red lines represent the estimations of the real sizes of the third COVID-19 epidemic wave in Qatar for the optimal value *β*_3_ = 5.308 in relationship (6): *V(t)=I(t)+R(t)* – solid; the dashed line represents the number of infectious persons multiplied by 10, i.e. *I* (*t*) × 10; the dotted line shows the derivative (*dV* / *dt*)×100 calculated with the use of formula (5). “Circles” and “stars” correspond to the accumulated numbers of cases registered during the period of time taken for SIR simulations (April 25 – May 8, 2021) and beyond this time period, respectively.

**Figure.**
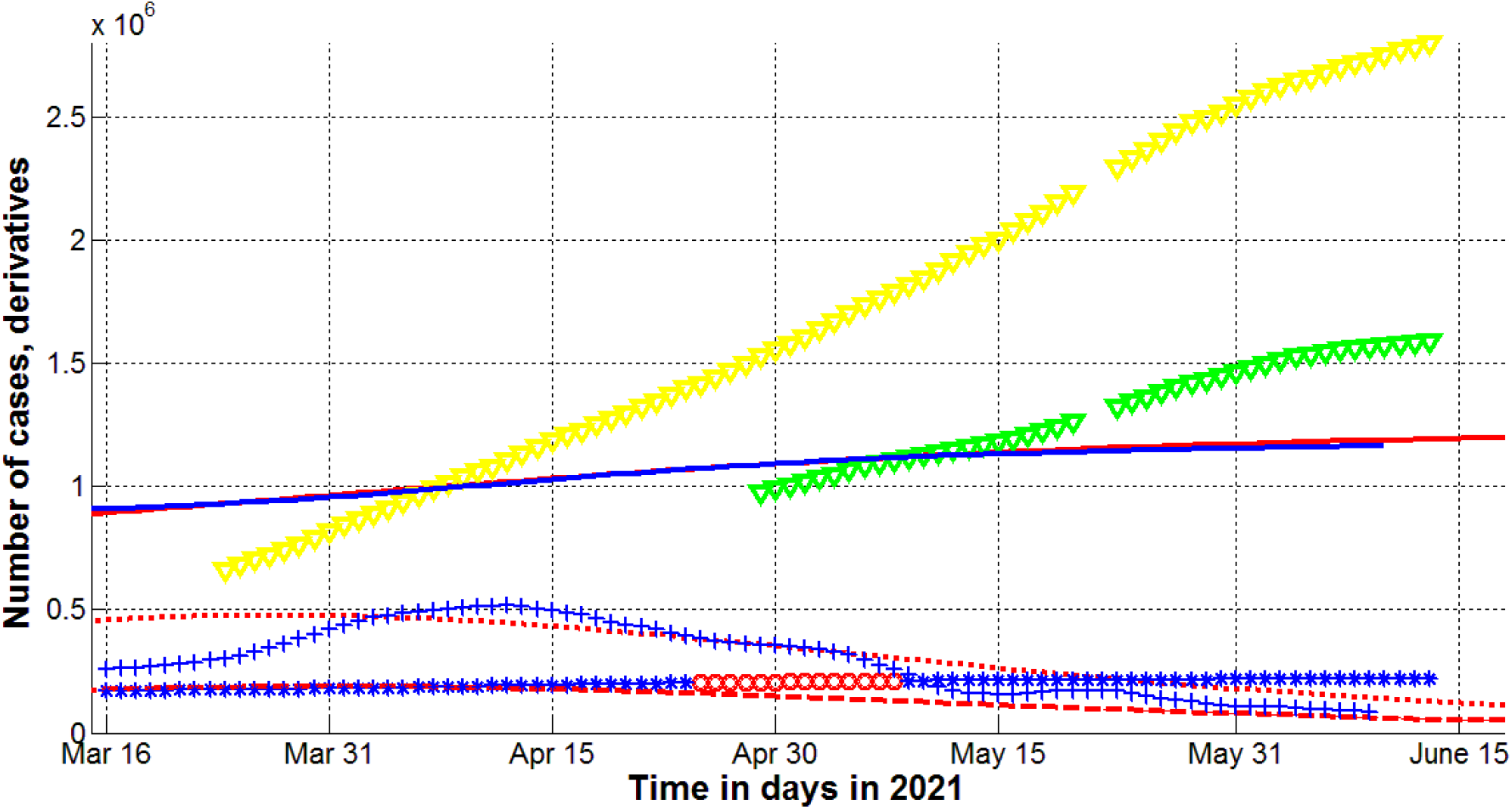
Real sizes of the third pandemic wave in Qatar at the calculated optimal value of the visibility coefficient *β*_3_ = 5.308. SIR simulations (red lines, [4]), numbers of vaccinations (yellow, Table 1) and vaccinated persons (green, Table 1). Numbers of victims *V(t)=I(t)+R(t)* – the red solid line; numbers of infected and spreading *I(t)* multiplied by 10 – dashed; derivative *dV/dt* (eq. (5)) multiplied by 100 – dotted, [4]. Markers show accumulated numbers of laboratory-confirmed cases *V*_*j*_ (from Table 1 and [4]) and derivatives. Red “circles” correspond to the accumulated numbers of cases taken in [4] for calculations of the third wave; blue “stars” – number of cases beyond this period. “Crosses” show the values of the first derivative (eq. (2)) multiplied by 100*β*_3_. The blue solid line represents smoothed accumulated number of laboratory-confirmed cases (1) multiplied by *β*_3_.

To illustrate the accuracy of calculations, the real accumulated number of laboratory confirmed cases *V*_*j*_ form Table 1 and Table 1 in [4] was smoothed with the use of (1) and multiplied by the “visibility” coefficient *β* = 5.308, calculated in [4]. The result is shown in the figure by the solid blue line and is in very good agreement with the theoretical estimation (4) obtained in [4] and shown by the solid red line. We have calculated also the first derivative (2) with the use of eq. (1). The corresponding values have been multiplied by 100*β* and shown by blue “crosses”. These values are in rather good agreement with the theoretical estimation of the real daily number of new cases (eq. (5), the dotted red line, [4]).

The epidemic dynamics in June 2021 shows that the averaged number of new cases (eqs. (1) and (2), blue “crosses”) started to deviate from the theoretical estimation (eq. (5), the dotted red line) obtained in [4] with the use of data set corresponding the time period April 25 – May 8, 2021. Probably it is a result of mass vaccination, since the number of vaccinated people exceeded the half of Qatar population in June 2021 (see green markers). It would be very interesting to follow the further dynamics of the pandemic in Qatar in order to assess the long-term impact of vaccination

Unfortunately, mathematical modeling cannot answer the question about possibility new epidemic waves after June 2021, because even strict quarantine measures does not prevent the emergence of new strains of coronavirus. The number of infectious persons in Qatar is still high. According to the estimation (shown by the red dashed line), the number of infectious persons was around 6,000 in mid-June 2021. This fact and the unfavorable course of the pandemic in India [12, 13] make the probability of the emergence of new coronavirus strains and new epidemic waves rather high. In the case of an optimistic scenario (without the emergence of new waves), the end of the epidemic in Qatar should be expected in March 2022 (see [4]). The actual number of cases may increase during this rather long period (from June 13, 2021) by 70,000 (the number of new registered cases will be about five times less). The government should closely monitor the average daily number of cases and its derivative (which can be calculated by formulas (1)-(3)) and immediately intensify quarantine measures in case of their increase.

## Data Availability

data is in the text

## Acknowledgements

The author is grateful to Oleksii Rodionov for his help in collecting and processing data.

## References

1. Ahmed E. Fahmya, Mohammed M. El-desoukya, Ahmed S.A. Mohamed. Epidemic Analysis of COVID-19 in Egypt, Qatar and Saudi Arabia using the Generalized SEIR Model. MedRxiv. Posted August 22, 2020. Doi: https://doi.org/10.1101/2020.08.19.20178129

2. Ryad Ghanam, Edward L. Boone, Abdel-Salam G. Abdel-Salam. SEIRD MODEL FOR QATAR COVID-19 OUTBREAK: A CASE STUDY. Posted 26 May 2020. arXiv:2005.12777v1

3. Nesteruk I., Benlagha N. PREDICTIONS OF COVID-19 PANDEMIC DYNAMICS IN UKRAINE AND QATAR BASED ON GENERALIZED SIR MODEL. Innov Biosyst Bioeng, 2021, vol. 5, no. 1, pp. 37–46. Doi: 10.20535/ibb.2021.5.1.228605 http://ibb.kpi.ua/article/view/228605

4. Nesteruk I. Impact of vaccination and undetected cases on the COVID-19 pandemic dynamics in Qatar in 2021. [Preprint] MedRxiv, June 2021. Doi: 10.1101/2021.05.27.21257929 https://medrxiv.org/cgi/content/short/2021.05.27.21257929v1

5. COVID-19 Data Repository by the Center for Systems Science and Engineering (CSSE) at Johns Hopkins University (JHU). https://github.com/owid/covid-19-data/tree/master/public/data

6. Nesteruk I. Visible and real sizes of new COVID-19 pandemic waves in Ukraine. Innov Biosyst Bioeng, 2021, vol. 5, no. 2, pp. 85–96. Doi: 10.20535/ibb.2021.5.2.230487 http://ibb.kpi.ua/article/view/230487

7. Nesteruk I. COVID-19 pandemic dynamics. Springer Nature, 2021, DOI: 10.1007/978-981-33-6416-5. https://link.springer.com/book/10.1007/978-981-33-6416-5

8. Nesteruk I. Detections and SIR simulations of the COVID-19 pandemic waves in Ukraine. Comput. Math. Biophys. 2021;9:46–65. https://doi.org/10.1515/cmb-2020-0117

9. Nesteruk I. Identification of the New Waves of the COVID-19 Pandemic. In book: COVID-19 Pandemic Dynamics, Springer Nature, 2021. Doi: 10.1007/978-981-33-6416-5_8. https://link.springer.com/chapter/10.1007/978-981-33-6416-5_8

10. Nesteruk I. General SIR Model and Its Exact Solution. In book: COVID-19 Pandemic Dynamics, Springer Nature, 2021. Doi: 10.1007/978-981-33-6416-5_9. https://link.springer.com/content/pdf/10.1007%2F978-981-33-6416-5_9.pdf

11. Nesteruk I. Procedures of Parameter Identification for the Waves of Epidemics. In book: COVID-19 Pandemic Dynamics, Springer Nature, 2021. Doi: 10.1007/978-981-33-6416-5_10. https://link.springer.com/chapter/10.1007%2F978-981-33-6416-5_10

12. Nesteruk I. The COVID-19 pandemic storm in India. [Preprint] medRxiv Posted May 08, 2021. Doi: https://doi.org/10.1101/2021.05.06.21256523

13. Nesteruk I. The COVID-19 pandemic storm in India subsides, but the calm is still far away. [Preprint] medRxiv June, 2021. Doi: https://doi.org/10.1101/2021.06.01.21258143

